# COVID-19 infection among health care workers: Experience in Base Hospital Wathupitiwala, Sri Lanka

**DOI:** 10.1101/2021.08.28.21262733

**Authors:** W.A.M.P. Samaranayake, G.P.C. Jayawardena, A.L.L. Roshan, M.A.M. Wijewardene, M.I. Siraj

**Author notes:** Correspondence: W.A.M.P.Samaranayake. No:57/17, Railway Station Road, Veyangoda.

## Abstract

Coronavirus disease 2019 (COVID-19) is a serious global health pandemic resulting in high mortality and morbidity. Frontline health care workers (HCWs) are at an increased risk of the acquisition of severe acute respiratory syndrome coronavirus-2 infection (SARS CoV-2) due to their close interaction with infected patients (1, 2). Also, HCWs can serve as reservoirs of SARS CoV-2 cross-transmission both in community and hospital settings (1). However, the extent of COVID-19 infection among HCWs in Sri Lanka is understudied.

**Objectives:** This study determined the incidence, demographic characteristics, and risk exposure behavior of HCWs who tested positive for SARS CoV-2 at Base Hospital Wathupitiwala. Furthermore, the rate of acquisition of SARS CoV-2 following COVISHIELD/ChAdOx1 nCoV-19 and Sinopharm /BBIBP-CorV vaccines in HCWs were studied.

**Methods:** A retrospective cross-sectional descriptive analysis was conducted from May 2021 to August 2021 for a total of 818 HCWs.

**Results:** Hundred and twenty-four HCWs (15.16%) were tested positive for COVID-19. The mean age of infected HCWs was 46.27 years and the majority were females (74.19%). Among all infected persons, 54 (43.55%) were nurses/midwives, 39 (31.45%) were clinical supportive staff and 12(9.68%) were medical officers. The number of infected HCWs rapidly escalated and a total of 64(51.61%) HCWs got an infection during August/2021. No source was identified in most of them (34.68%) followed by community acquisition (33.87%). Thirty-five HCWs (28.23%) had acquired infection during a hospital setting or had a high-risk exposure in recent history. Among hospital-related infections, 37.91% of HCWs had shared meals or shared sleeping rooms with an infected workmate. The majority of the HCWs were tested by the infection control unit as symptomatic screening (70.16%) followed by contact tracing (20.16%). Fifty-six (45.16%) HCWs had a history of single or multiple comorbidities. The vast majority of HCWs (95.97%) presented as mild to asymptomatic disease that followed an uneventful recovery. Body aches, headache, fever, and sore throat were the most commonly reported symptoms among them. Among the five HCWs required therapeutic oxygen supplementation, two unvaccinated HCWs succumbed to the infection. The rate of breakthrough infection among HCWs was 8.93%. The acquisition of disease was significantly higher among unvaccinated HCWs than partially (p<0.0001) or fully vaccinated (p<0.0001) HCWs with either type of vaccine.

**Conclusions:** Protecting HCWs remains a challenge in resource-poor settings. The risk of infection fueled by very contagious circulating variants is continuously high even though vaccination has shown clear benefits in preventing mortality and severe infection. Therefore, all healthcare workers should be vaccinated while ensuring continuous infection control measures in the hospital setting.

## Introduction

The coronavirus disease 2019 (COVID-19) pandemic caused by severe acute respiratory syndrome coronavirus 2 (SARS-CoV-2) is the most fearsome challenge to humanity in recent decades. COVID-19 has overwhelmed hospital capacity and jeopardized the health care system in most countries (1, 2).

Health care workers (HCWs) are a particularly high-risk group due to their close interaction with infected persons (1, 3). A systemic review of 237 published studies that evaluated epidemiology and risk factors associated with SARS-CoV-2 infection in HCWs found a wide variation in seroprevalence ranging from 0.3% to 40%, with numbers probably underestimated (3,4). However, higher infection rates among HCWs were associated with unprotected exposures to COVID-19 patients or infected workmates while performing certain high-risk procedures or direct physical contact with the patients or their body secretions (4).

Sri Lanka reported the first case of COVID-19 on 27th January 2020 and thereafter several waves occurred at different intensities, mainly in the Western province, such as the Navy officers’ and Minuwangoda clusters. Subsequently, the fish market-related cluster was linked to almost all the districts, followed by a highly intensified New Year-related outbreak in April that resulted in a substantial degree of community spread (5). The recent rapid surge of infective cases and deaths that occurred in August 2021 had dramatically crippled the health system due to the widespread delta (B.1.617.2) variant (5, 6) across the country. However, the true extent of the disease among HCWs has been difficult to determine in Sri Lanka, yet PCR testing from nasopharyngeal swabs is challenging with limited resources. Furthermore, the COVID-19 infection in Sri Lankan HCWs is understudied.

Sri Lanka’s COVID-19 response is not very different from the transmission-mitigation strategies practiced worldwide (5). However, it is believed that a large-scale vaccination is the most effective public health strategy to mitigate the COVID-19 pandemic to rapidly reduce hospitalization, severe outcomes, and deaths while achieving herd immunity (4). In the current context, the government of Sri Lanka commenced the national COVID-19 vaccination program following its emergency approval of Oxford-AstraZeneca/ChAdOx1 nCoV-19, Sinopharm/BBIBP-Cor V, and Sputnik-V/Gam COVID-Vac and Pfizer-BioNTech/tozinameran. In addition, Jeewandara et al., 2021 recently showed a high seroconversion rate following two doses of Sinopharm/BBIBP-Cor V in his study (6). However, an in-depth analysis of the clinical data and real-world impacts of COVID-19 vaccines, along with immunogenicity data, is essential to comprehend the real picture in Sri Lanka. Yet, there are no published studies to assess the real-world effects of different COVID-19 vaccines among HCWs in Sri Lanka. Moreover, it is unknown whether the clinical characteristics, the trend of the disease, and outcomes of COVID 19 infection in HCWs are different from those of the general population.

Therefore, this study was designed to determine the incidence of COVID-19 infection and the epidemiology and demographic and high-risk exposure behavior characteristics of HCWs at Base Hospital Wathupitiwala. Also, this study determined the rate of breakthrough infection following the COVISHIELD/ChAdOx1 nCoV-19 and Sinopharm/BBIBP-Cor V vaccines.

## Materials and Methods

This was a retrospective cross-sectional descriptive analysis of all HCWs who tested positive for COVID-19 during the period from May 2020 to August 2021 at Base Hospital Wathupitiwala, Sri Lanka. This institute has a 600-bed capacity and employs a total of 818 health care workers. The HCWs were tested if they reported symptoms or those with a history of any significant exposure with a confirmed/suspected case or as a part of hospital random screening. The testing was performed using a deep nasopharyngeal swab through a validated Real-time reverse transcription-polymerase chain reaction (rRT-PCR) at the reference laboratory or Rapid Antigen test (STANDARD Q COVID-19 Ag test, SD Biosensor, Republic of Korea) by a trained medical laboratory technician at Base Hospital Wathupitiwala (7). All cases were notified to the infection control unit headed by a Consultant Microbiologist and investigated by a trained medical officer to determine the level of risk and type of acquisition. The risk assessment was conducted according to the circular issued by the Ministry of Health and Indigenous Medicine (8), Sri Lanka, and the HCWs guidelines of the Center for Disease Control and Prevention (CDC) (9). Case-based decisions were made with the help of the COVID operational cell committee that consists of a panel of experts at Base Hospital Wathupitiwala. All the data related to the epidemiology, social demography and follow-up communication of HCWs were recorded.

### Staff vaccination program

The national COVID-19 vaccination program began vaccinating HCWs against SARS-CoV-2 on 30th January 2021, initially using the COVISHIELD/ChAdOx1 nCoV-19 vaccine (two doses given three months apart) and subsequently with the Sinopharm/BBIBP-Cor V vaccine (two doses given 4 weeks apart) on 07th June 2021. This was made available to all the HCWs at Base Hospital Wathupitiwala on an ongoing basis. Data were extracted from the institutional vaccine registry for the study.

### Definitions

A health care worker was defined as all paid and unpaid people serving in a healthcare setting who have the potential for direct or indirect exposure to patients or infectious materials (9).

A significant exposure was considered as face-to-face contact with a person with COVID-19 within 6 feet for more than 15 minutes or any unprotected direct physical contact with a person with COVID-19 or infectious secretions from COVID-19 (9).

The infection was considered community-acquired if the HCW reported contact with a confirmed COVID-19 case in the community setting. The infection was considered hospital-acquired if the HCW reported exposure to a confirmed or suspected COVID-19 case in the hospital and had not had any contact with asymptomatic or confirmed COVID-19 case in the community or been exposed as per infection control unit records. HCWs with no exposure to symptomatic or suspected/confirmed cases in the hospital or the community were recorded as having an unidentified source (10, 11).

### Breakthrough infection

The detection of SARS CoV-2RNA or antigen in a respiratory specimen collected from a person more than 14 days after they have completed all recommended doses of Food and Drug Administration (FDA) authorized COVID 19 vaccine(9)

Ethic and review approval for this study was obtained from the Ethics and review committee at Lady Ridgway Hospital Colombo (LRH/DA/29/2020).

## Statistical Analysis

Data were entered into an SPSS software (version 17) and descriptive analysis was conducted. The number of infected HCWs was tabulated against the type and dose of the COVID-19 vaccine and a chi-square test was used to assess the difference between groups. A p-value of less than 0.05 was considered significant.

## Results

A total of 818 HCWs were included in the study. Nine hundred and fifty rRT-PCR tests and 810 Rapid Antigen Tests were performed between May/2020 and August/2021. A total of 124 (15.16%) HCWs were found infected. Figure 1 summarizes the spread of infected HCWs according to the study period, and numbers escalated rapidly during the month of August. Table 1 describes the epidemiology and demographic characteristics of COVID-19 infected HCWs at Base Hospital Wathupitiwala.

**Figure 1:**
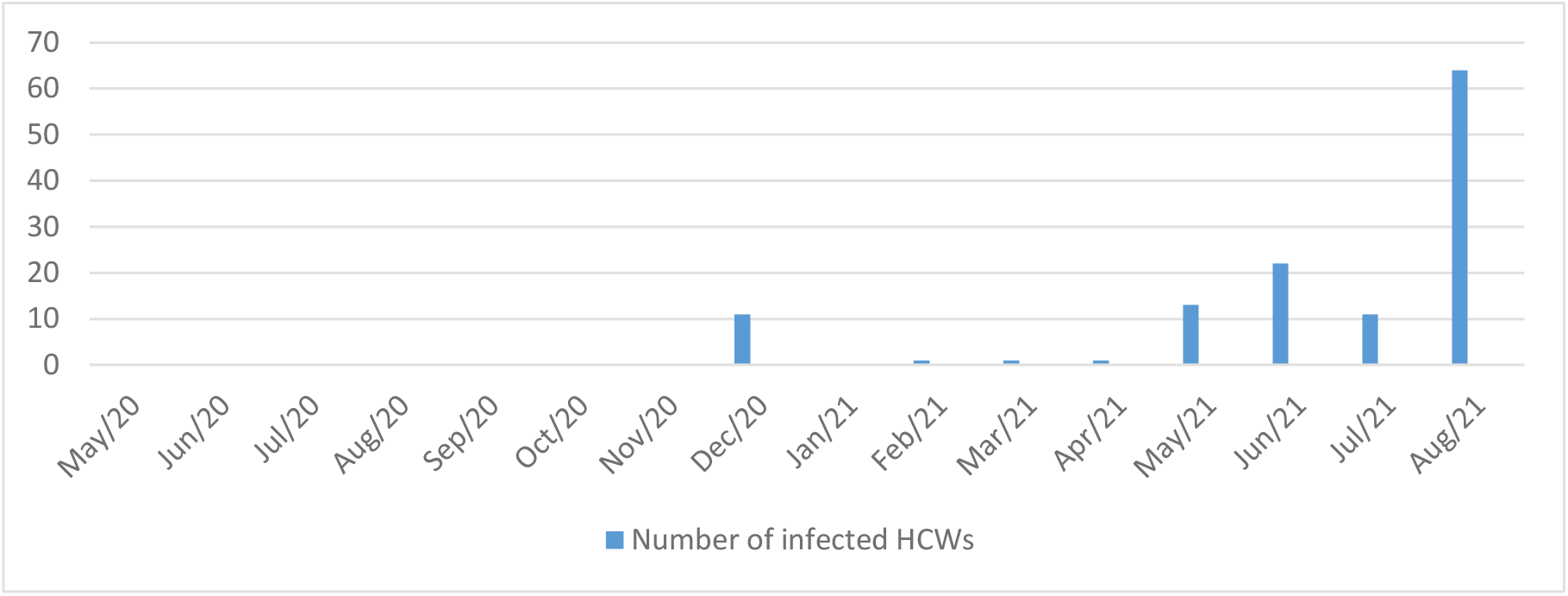
Number of infected health care workers during study period. Period(X): month /year, Number (Y): Number of infected health care workers

**Table 1:**
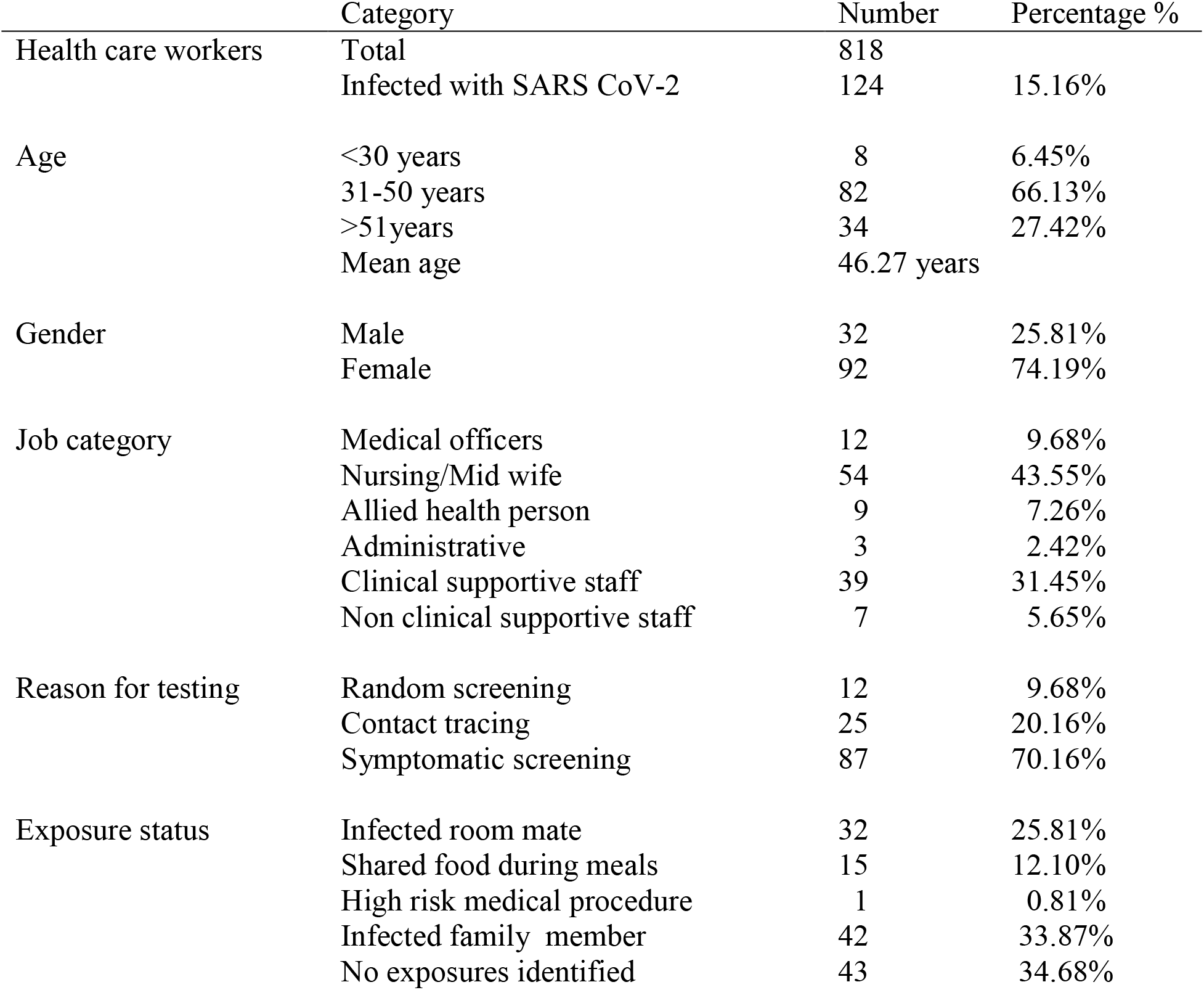
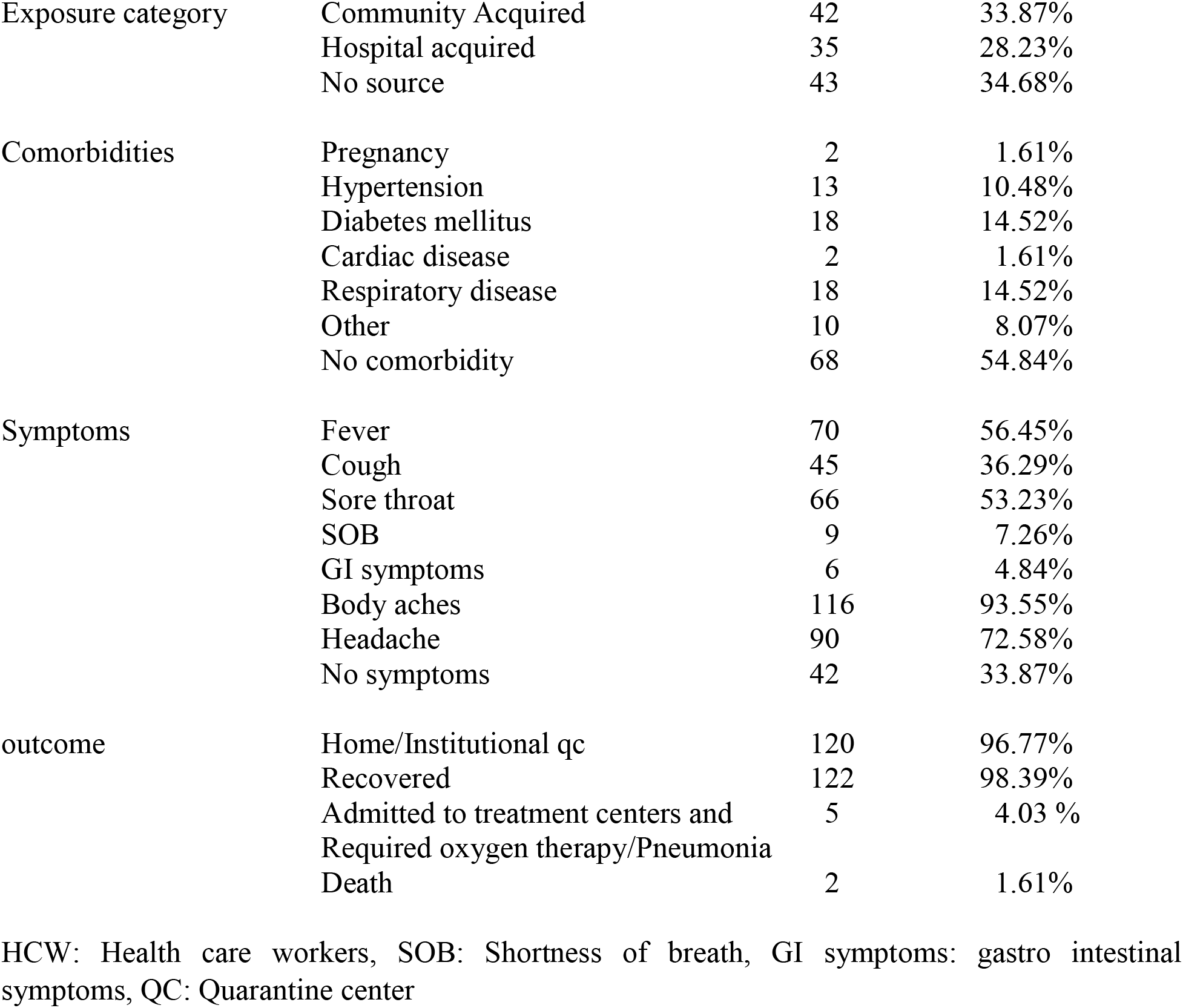
Demographic characteristics of HCWs who tested positive for SARS CoV-2 at Base Hospital Wathupitiwala.

Among all infected HCWs, hundred and nineteen (95.96%) developed mild to asymptomatic COVID-19 and followed an uneventful recovery. Body aches, headache were most commonly reported symptoms while diarrhea and shortness of breath were less commonly reported, as shown in figure 2. Table 2 shows how HCWs acquired the infection following vaccination. Among all, seventy-one (8.93%) had breakthrough infections while seventeen (2.13%) acquired infection following a single dose of vaccination. No HCW developed a severe breakthrough infection during this period. The association of infected HCWs was not significant between the two groups of COVISHIELD/ChAdOx1 nCoV-19 and Sinopharm /BBIBP-CorV vaccines irrespective of the doses received. However. The acquisition of disease was significantly higher among unvaccinated HCWs than partially (p<0.0001) or fully vaccinated (p<0.0001) HCWs with both types of vaccines.

**Figure 2:**
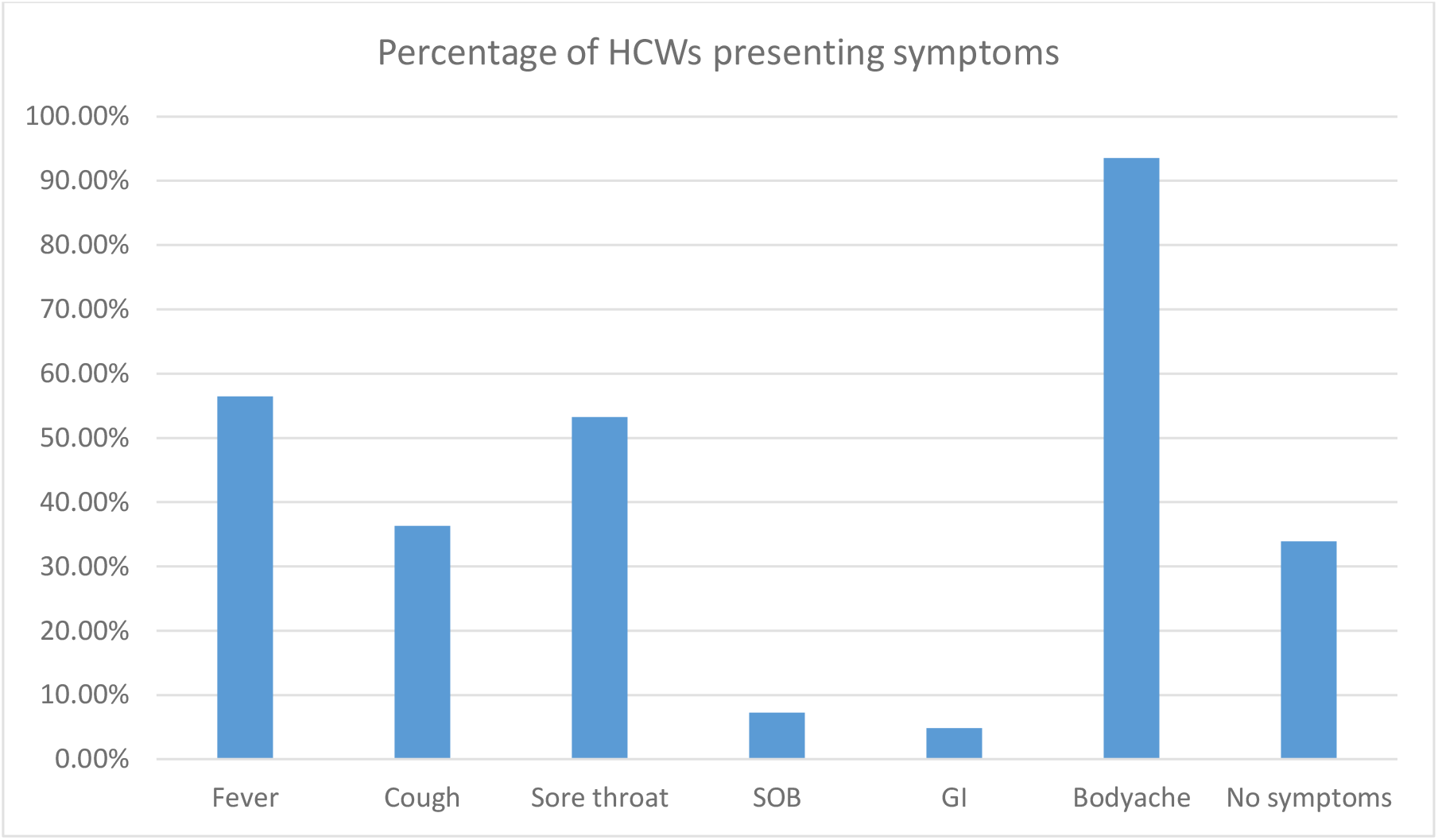
Percentages of HCWs presenting symptoms on diagnosis Presenting symptoms(x), Percentage of HCWs presenting symptoms (Y), SOB: Shortness of breath, GI: Gastro intestinal symptoms, HCW: Health care worker

**Table 2:** Occurrence of COVID-19 infection among health care workers after the COVID-19 vaccination at Base Hospital Wathupitiwala.

|  | Un vaccinated<br>/Percentage | Sinopharm /BBIBP-Cor V |  | COVISHIELD/ChAdOx1<br>nCoV-19 |  |
| --- | --- | --- | --- | --- | --- |
|  |  | Partially<br>vaccinated | Fully<br>vaccinated | Partially<br>vaccinated | Fully<br>vaccinated |
| HCWs number | 18 | 129 | 129 | 671 | 666 |
| Total infected HCWs | 4(22.22%) | 15(11.63%) | 14(10.85%) | 2(0.30%) | 57(8.56%) |
| Severe infections | 0 | 2(1.55%) | 0 | 1 | 0 |
| Total deaths | 2(11.11%) | 0 | 0 | 0 | 0 |

HCWs: Health care worker, Partially vaccinated: received single dose of vaccine, Fully vaccinated: received two doses of vaccination, COVID-19: Coronavirus disease 2019.

## Discussion

The COVID-19 situation is continuously a global challenge due to high levels of transmission dynamics driven by four major factors, such as virus variants of concern, increased social mixing and mobility of people, inappropriate use of public health and social measures, and vaccine inequity (1, 2, 4). Understanding of SARS CoV-2 transmission risk in the health care setting is critical for the resilience of the health system to successfully combat such a major pandemic (4). However, despite all efforts to protect HCWs, some exposures are inevitable.

According to our knowledge, this is the first published study of infected HCWs to assess the epidemiology, demography, risk exposure behaviors, and breakthrough infections rate in Sri Lanka. Our study shows that the high rate of acquisition of SARS-CoV-2 infection among young HCWs which is in line with global data (2, 3). It is noteworthy that the first infected HCW was detected at Base Hospital Wathupitiwala in December in the background of the Minuwangoda cluster. The small peak of infection in May and June 2021 has coincided with the New Year festival, which was an important cultural event characterized by large social gatherings and family visits. Thereafter, a dramatic rise of infection after July 2021 has overwhelmed the hospital services probably due to the prevailing Delta (B.1.617.2) variant across the Country. Half of the study population acquired the infection during the month of August. Overall, infection trends in incidence among HCWs at Base Hospital Wathupitiwala followed parallel to the general population in Sri Lanka. A similar phenomenon was observed in a large systemic review of 152888 COVID-19 infections in HCWs around the world (2, 3,).

Females got infections more commonly while nursing officers and midwives were at high risk of developing the disease, which is similar to global data (2, 3). Most of the infected HCWs (96.96%) developed mild to asymptomatic disease. A significant proportion of HCWs had nonspecific symptoms such as body aches, headache, fever, and sore throat. We believe that this may create the perfect environment for silent transmission both in the hospital and community contexts if preventive methods are not implemented (2).

It appears that the rate of community acquisition of SARS CoV-2 is slightly greater than that of hospital acquisition in our setting, although the majority had not identified any link to the source. This has caused increased pressure on the healthcare system at Base Hospital Wathupitiwala. Among hospital-related infections, the majority of HCWs acquired infections as a result of contact with another infected workmate, particularly during “rest” times, as the HCWs were not compliant with social distancing and universal masking while eating and sleeping in an enclosed environment. The data from the COVID-19 pandemic shows that appropriate use of personal protective equipment (PPE), and re-organization of the workflow process would reduce the risk of transmission of SARS CoV-2 to HCWs (12,13,14). However, our study was underpowered to assess other risk factors, such as the inadequate provision of PPE/Alcohol hand rub and work overload. In addition, we have observed in-built colonial building architecture with overcrowded small staff restrooms with inadequate ventilation as major challenges in implementing infection control practices in this institution.

Since the start of the COVID-19 vaccination program, the development of new infections has been reported worldwide following one or both doses of vaccine (15). Our finding shows that there is no difference of breakthrough infection in HCWs between two groups, Oxford-AstraZeneca (ChAdOx1 nCoV-19) and Sinopharm, /BBIBP-Cor V. However, acquisition of infection was significantly higher in an unvaccinated group compared to both the partial or fully vaccinated categories. Our study proves that the national COVID-19 vaccination program has prevented severe disease with zero mortality among HCWs at Base Hospital Wathupitiwala.

The rate of breakthrough infections at Base Hospital Wathupitiwala, is above the risk reported in the clinical trial of Oxford-AstraZeneca/ChAdOx1 nCoV-19 and Sinopharm/BBIBP-Cor V vaccines. These higher rates could be due to differences in demographic characteristics and health status profiles (16, 17). In addition, the alpha variant (B.1.1.7) was dominant in the early period from May to April 2021 whereas the highly infectious delta variant (B1.617.2) was predominant after July 2021 in Sri Lanka (5, 6). Moreover, the majority of our study participants had mild to asymptomatic disease. More conclusive studies are needed to assess how COVID - 19 vaccines can substantially reduce the transmission risk among vaccinated HCWs. In the context of a higher rate of breakthrough infections, the need for an additional vaccine dose for front-line health care workers in Sri Lanka should be carefully be evaluated along with data on weaning immunity, vaccine variant of concern, and type of vaccine given. Therefore, we believe that our study underscores the critical importance of continuing public health mitigation strategies, even in an environment with a high incidence of vaccination, until herd immunity is reached at large.

All five HCWs who developed severe disease that required supplemented oxygen had single or multiple comorbidities such as Diabetes mellitus, Bronchial Asthma and dyslipidemia. One HCW had multiple myeloma in remission. Among them, two HCWs had the disease a few days after the first jab of Sinopharm/BBIBP-Cor V which can be considered as almost non vaccinated individuals. The exact immunological mechanism causing progression to severe disease is not yet well understood (18). One partially vaccinated HCW got the infection four months after the first jab of the COVISHIELD/ChAdOx1 nCoV-19 vaccine. This could be explained by the possibility of a weaning effect of antibody response. The death rate (1.61%) of infected HCWs at Base Hospital Wathupitiwala is considerably low and no vaccinated HCW had died during that period. In contrast, general data indicates that 7% of the higher death toll in Sri Lanka is probably due to the underreporting of a total number of cases. A recent systematic review and meta-analysis revealed that a pooled prevalence of severe COVID-19 was 5% among HCWs, while 0.5% of the infected HCWs had died because of complications of the disease (2, 3).

This study has several limitations. The actual number of infected HCW might be less due to underreporting of symptoms like a high proportion of patients with COVID-19 remained asymptomatic. The findings of our data related to vaccines should be interpreted with caution as those observations were made for only a short duration. Analysis of large-scale clinical data for a long duration with a randomized double-blind placebo control strategy will better reflect the real-world impacts of COVID-19 vaccines among HCWs.

## Conclusion

Protecting HCWs remains a challenge in resource-poor settings. The risk of infection fueled by very contagious circulating variants is continuously high even though vaccination has shown clear benefits in preventing mortality and severe infection. Therefore, all healthcare workers should be vaccinated while ensuring continuous infection control measures in the hospital setting.

## Data Availability

All data is available in the paper. The data-set generated during the current study are available from the corresponding author on reasonable request.

## Acknowledgements

We would like to acknowledge the infection control nurses for their support in collecting data and help in analyzing hospital related exposure incidents and Medical Laboratory technologists at Base Hospital Wathupitiwala and National Cancer Institute at Maharagama for performing Rapid Antigen test and r RT-PCR for SARS CoV-2 infection. We thanks for Dr. Aravinda Wickramasinghe, Registrar in Community Medicine for helping in statistical calculations.

